# Elevated serum IgM levels indicate poor outcome in patients with coronavirus disease 2019 pneumonia: A retrospective case-control study

**DOI:** 10.1101/2020.03.22.20041285

**Authors:** Zhihua Wang, Huijun Li, Jingjing Li, Chunguang Yang, Xiaolin Guo, Zhiquan Hu, Zhiqiang Chen, Shaogang Wang, Jihong Liu

**Affiliations:** Department of Urology, Tongji Hospital, Tongji Medical College, Huazhong University of Science and Technology, Wuhan, China; Department of Clinical Laboratory, Tongji Hospital, Tongji Medical College, Huazhong University of Science and Technology, Wuhan, China; Department of Clinical Laboratory, Union Hospital, Tongji Medical College, Huazhong University of Science and Technology, Wuhan, China

**Keywords:** 2019-nCoV, SARS-CoV-2, COVID-19, Coronavirus, Immunoglobulin

## Abstract

**Background:** The coronavirus disease 2019 (COVID-19) pneumonia outbreak began in Wuhan and pandemics tend to occur. Although SARS-CoV-2-specific immunoglobulins have been detected in serum of COVID-19 patients, their dynamics and association with outcomes have not been characterized.

**Methods:** A total of 116 hospitalized patients with confirmed COVID-19 pneumonia and SARS-CoV-2-specific immunoglobulins tested in Tongji hospital were retrospectively investigated. Clinical, laboratory, radiological characteristics and outcomes data were compared between mild-moderate group and died group. Further, a paired case-control study was conducted where each deceased case was matched to three mild-moderate patients of similar age.

**Findings:** Among 116 subjects included, 101 mild-moderate patients survived and 15 cases died. SARS-CoV-2-specific IgM levels peaked in forth week after onset of COVID-19 pneumonia, while serum IgG levels increased over 8 weeks. Serum IgM levels were higher in deceased patients than mild-moderate patients (P = 0.024), but not IgG. Serum IgM levels were negatively correlated with clinical outcome, eosinophil count and albumin levels (r = −0.269, P = 0.003; r = −0.188, P = 0.043; and r = −0.198, P = 0.033, resp.). The area under the ROC curve (AUC) for IgM antibody was 0.681 (95% CI: 0.517-0.845, P = 0.024). In case-control study paired by age, serum IgM was higher in deceased patients than mild-moderate patients (P = 0.019), positively correlated with leucocyte count (r = 0.260, P = 0.045), while negatively correlated with clinical outcome and albumin levels (r = −0.337, P = 0.008; r = −0.265, P = 0.041). AUC for IgM levels was 0.704 (95% CI: 0.534-0.873, P = 0.019).

**Interpretation:** These results indicate that dynamics of SARS-CoV-2 specific IgM and IgG antibodies was similar with that of SARS-CoV, while elevated serum IgM levels indicate poor outcome in patients with COVID-19 pneumonia.

## Introduction

Coronavirus disease 2019 (COVID-19) has spread rapidly worldwide since its discovery in December 2019^1,2^. As of March 14, 2020, over five thousand death from 2019 novel coronavirus (SARS-CoV-2) infection have been reported in 43 countries^3^. Effective strategies to treat this disease are urgently needed, but no drugs have been proven to be effective for the treatment of COVID-19 pneumonia ^4^. Antibodies are key components in the immune responses to viral infections^5,6^. Raised SARS-CoV-2-specific immunoglobulin M (IgM) and immunoglobulin G (IgG) have been detected in serum of COVID-19 patients^7^. However, less information is available about their dynamics and role in COVID-19 pneumonia.

Therefore, 116 hospitalized patients with COVID-19 pneumonia and with SARS-CoV-2 specific serum IgM and IgG detected were retrospectively selected between February 23, 2020, and March 11, 2020. The dynamics and role of serum IgM and IgG were investigated in this study.

## Methods

### Study design and participants

A total of 116 hospitalized patients with confirmed COVID-19 pneumonia and with serum IgM and IgG against SARS-CoV-2 examined between February 23, 2020, and March 11, 2020 in Tongji Hospital were retrospectively investigated. In brief, the patients in mild-moderate group described here were all probable hospitalized subjects in four wards of Tongji hospital with (1) serum IgM and IgG against SARS-CoV-2 examined, (2) chest radiographic evidence of pneumonia, (3) positive throat swab nucleic acid test by real-time RT-PCR methods or ≥ a fourfold increase in specific antibodies by the chemiluminescence kit. Patients with severe pneumonia^8^ or discharged from hospital in 24 hours were excluded from mild-moderate group. Patients in died group were all probable hospitalized cases who died between February 23, 2020, and March 11, 2020 with serum IgM and IgG against SARS-CoV-2 detected. Similarly, Patients who died in 24 hours after hospitalization were excluded from this study. This study was reviewed and approved by the Medical Ethical Committee of Tongji Hospital of Huazhong University of Science and Technology (IRB ID:TJ-IRB20200343).

### Data collection

Information collection was accomplished through our hospital’s electronic medical record system. Clinical, laboratory, radiological parameters and clinical outcomes were obtained with standardised forms for all subjects involved. Two researchers independently reviewed the data.

### Antibody measurement

Serum IgM and IgG antibodies were detected by the chemiluminescence kit (iFlash-SARS-CoV-2 IgM, iFlash-SARS-CoV-2 IgG) and iFlash 3000 CLIA System supplied by Yhlo Biotech Co. LTD (Shenzhen, China), of which the chemiluminescence kit had completed the EU CE certification. Threshold of 10 AU/ml was used for both IgM and IgG as manufacturer recommended.

### Statistical analysis

The statistical software SPSS 23.0 was used in this study. Categorical variables were described as frequency rates and percentages, and continuous variables were described using mean with interquartile range (IQR). When the data were normal, continuous variables were analysed by t tests, otherwise using the Mann-Whitney test. Proportions for categorical variables were compared using the χ2 test. Receiver operating characteristic (ROC) curves were used to evaluate IgM and IgG as potential predictors for clinical outcome. Correlation analysis was evaluated by the Pearson test. All tests were 2-sided, and P < 0.05 was considered statistically significant.

## Results

### The characterization of patients

Among 116 subjects included, 101 mild-moderate patients survived and 15 cases died, including 65 males and 51 females (Table 1). Of the 116 patients, 77 (66%) had one or more comorbidities. Hypertension (44 [38%]), diabetes (23 [20%]), cardiovascular disease (10 [9%]), COPD (6 [5%]), malignancy (7 [6%]) and chronic renal disease (6 [5%]) were the most common coexisting conditions. The chief complaints were fever (90 [76%] of 116 patients), cough (17 [15%]), and other uncommon symptoms (Table 1). Of 116 patients, 95 (82%) patients were bilateral involvement evaluated on the chest CT or X-ray images. The mean hospitalized duration was 19.1 (6.0-29.0) days. People who died were significantly older (68.9 years [IQR 57.0-79.0] vs 60.0 years [IQR 50.0-69.0]; P = 0.019), more men included (12 [80%] vs 53 [52%]; P = 0.045) and more likely to have comorbidities (14 [93%] vs 63 [62%]; P = 0.018).

**Table 1:** Baseline characteristics of patients included in the study.

|  | **Total (n=116)** | **Mild-Moderate**<br>**(n=101)** | **Died**<br>**(n=15)** | **P value** |
| --- | --- | --- | --- | --- |
| **Age, years** | 61.1(51.0-69.0) | 60.0(50.0-69.0) | 68.9(57.0-79.0) | 0.019 |
| **Sex** |  |  |  | 0.045 |
| Male | 65(56%) | 53(52%) | 12(80%) |  |
| Female | 51(44%) | 48 (48%) | 3(20%) |  |
| **Any comorbidities** | 77(66%) | 63(62%) | 14(93%) | 0.018 |
| Hypertension | 44(38%) | 37(37%) | 7(47%) | 0.455 |
| Diabetes | 23(20%) | 20(20%) | 3(20%) | 0.986 |
| Cardiovascular disease | 10(9%) | 7(7%) | 3(20%) | 0.092 |
| COPD | 6(5%) | 5(5%) | 1(7%) | 0.779 |
| Malignancy | 7(6%) | 3(3%) | 4(27%) | <0.001 |
| Chronic renal disease | 6(5%) | 5(5%) | 1(7%) | 0.779 |
| Others | 19(16%) | 16(16%) | 3(20%) | 0.685 |
| **Chief complaint** |  |  |  |  |
| Fever | 90(76%) | 79(78%) | 11(73%) | 0.606 |
| Cough | 17(15%) | 13(13%) | 4(27%) | 0.121 |
| Dyspnea | 3(3%) | 3(3%) | 0(0) | 0.499 |
| Others | 6(5%) | 6(6%) | 0(0) | 0.332 |
| **SARS-CoV-2 RNA** |  |  |  | 0.838 |
| Positive | 95(82%) | 83(82%) | 12(80%) |  |
| Negative | 21(18%) | 18(18%) | 3(20%) |  |
| **Bilateral involvement** |  |  |  | 0.259 |
| Yes | 108(93%) | 93(92%) | 15(100%) |  |
| No | 8(7%) | 8(8%) | 0(0) |  |
| **Hospital stay** | 19.1(6.0-29.0) | 18.6(3.5-29.0) | 22.6(21.0-29.0) | 0.105 |

### Laboratory parameters in mild-moderate and deceased patients

The blood counts of patients on admission showed significantly decrease in lymphocytes, especially in died group than in mild-moderate patients (Table 2). Prothrombin time and D-dimer level on admission were higher in deceased patients (PT 15.5 s [14.6-16.6]; D-dimer 5.9 μg/L [0.8-6.8]) than mild-moderate patients (PT 14.0 s [13.3-14.4], P = 0.003; D-dimer 2.3 μg/L [0.3-1.9], P = 0.011). Blood urea nitrogen and creatinine level on admission were higher in deceased patients (BUN 10.9 mmol/L [7.5-11.9]; Creatinine 159.9 μmol/L [80.0-122.0]) than mild-moderate patients (BUN 5.7 mmol/L [3.9-6.5], P < 0.001; Creatinine 88.2 μmol/L [57.0-81.0], P < 0.001). Serum albumin level was lower in deceased patients (albumin 31.7 g/L [27.4-37.3]) than mild-moderate patients (albumin 36.0 g/L [31.7-40.4], P = 0.024). No significant differences existed in serum IL6 levels between deceased patients (15 patients included, 83.4 pg/mL [14.0-65.2]) and mild-moderate patients (91 patients included, 36.5 pg/mL [1.7-25.3], P = 0.093).

**Table 2:** Laboratory findings of patients with COVID-19 pneumonia.

|  | **Total (n=116)** | **Mild-Moderate**<br>**(n=101)** | **Died**<br>**(n=15)** | **P value** |
| --- | --- | --- | --- | --- |
| Hemoglobin, g/L | 126.0(115.3-138.0) | 124.9(115.5-137.0) | 133.1(114.0-149.0) | 0.123 |
| Alanine aminotransferase, U/L | 29.8(16.0-37.0) | 29.6(16.0-37.0) | 30.5(18.0-39.0) | 0.884 |
| Aspartate aminotransferase, U/L | 31.9(19.3-38.8) | 31.4(19.0-37.0) | 35.3(21.0-45.0) | 0.501 |
| Albumin, g/L | 35.5(30.7-40.3) | 36.0(31.7-40.4) | 31.7(27.4-37.3) | 0.017 |
| Total bilirubin, μmol/L | 11.0(6.6-14.1) | 10.7(6.3-14.2) | 13.3(9.2-13.9) | 0.119 |
| Lactate dehydrogenase, U/L | 312.5(198.8-367.5) | 299.5(194.5-330.0) | 399.9(262.0-524.0) | 0.055 |
| Blood urea nitrogen, mmol/L | 6.3(3.9-7.1) | 5.7(3.9-6.5) | 10.9(7.5-11.9) | <0.001 |
| Creatinine, μmol/L | 97.5(59.0-91.8) | 88.2(57.0-81.0) | 159.9(80.0-122.0) | <0.001 |
| Prothrombin time, second | 14.2(13.3-14.6) | 14.0(13.3-14.4) | 15.5(14.6-16.6) | 0.003 |
| D-dimer, μg/mL | 1.9(0.3-2.2) | 2.3(0.3-1.9) | 5.9(0.8-6.8) | 0.011 |
| IgM, AU/ml | 110.0(13.3-104.9) | 88.6(12.9-88.1) | 253.8(29.2-127.3) | 0.024 |
| IgG, AU/ml | 201.4(149.5-239.8) | 197.1(149.2-225.3) | 230.7(157.3-295.5) | 0.202 |
| Antibody detection after onset, day | 31.6(27.0-38.0) | 31.9(27.0-39.5) | 30.1(26.0-32.0) | 0.228 |
| Leucocytes, $\times 10^9$ per L | 7.4(4.8-8.1) | 7.1(4.8-7.7) | 9.4(7.3-11.5) | 0.052 |
| Neutrophils, $\times 10^9$ per L | 5.6(2.9-6.3) | 5.2(2.8-5.7) | 8.2(6.2-10.0) | 0.014 |
| Lymphocytes, $\times 10^9$ per L | 1.1(0.7-1.5) | 1.2(0.7-1.6) | 0.7(0.5-0.9) | 0.002 |
| Monocytes, $\times 10^9$ per L | 0.5(0.3-0.6) | 0.5(0.4-0.6) | 0.5(0.3-0.7) | 0.924 |
| Eosinophils, $\times 10^9$ per L | 0.1(0.0-0.1) | 0.1(0.0-0.1) | 0.0(0.0-0.0) | 0.001 |
| Basophils, $\times 10^9$ per L | 0(0.0-0.0) | 0.0(0.0-0.0) | 0.0(0.0-0.0) | 0.675 |
| Platelets, $\times 10^9$ per L | 230.9(158.5-300.8) | 233.6(165.0-301.5) | 212.9(113.0-295.0) | 0.172 |

### IgM levels were associated with clinical outcome

Serum IgM levels were higher in deceased patients (IgM 253.8 AU/ml [29.2-127.3]) than mild-moderate patients (IgM 88.6 AU/ml [12.9-88.1], P = 0.024), but not IgG (230.7 AU/ml [IQR 157.3-295.5] vs 197.1 AU/ml [IQR 149.2-225.3]; P =0.202; Table 2, Figure 1A, B). We subsequently explored the correlations between IgM levels and clinical outcome, serum IgG levels, blood counts, or other involved parameters. Interestingly, serum IgM levels were positively correlated with IgG levels (r = 0.251, P = 0.006), while negatively correlated with clinical outcome, eosinophil count and albumin levels (r = −0.269, P = 0.003; r = −0.188, P = 0.043; and r = −0.198, P = 0.033, resp.). Meanwhile, no significant correlations were found between IgM levels and sex, comorbidities, leucocyte count, neutrophil count, lymphocyte count, blood urea nitrogen levels, creatinine levels, PT, D-dimer levels, IL6 levels or duration between onset and antibody detection in patients with COVID-19 pneumonia (r = −0.139, P = 0.136; r = −0.002, P = 0.979; r = 0.165, P = 0.076; r = 0.171, P = 0.067; r = −0.052, P = 0.582; r = 0.005, P = 0.954; r = −0.081, P = 0.390; r = 0.127, P = 0.175; r = 0.087, P = 0.354; r = 0.121, P = 0.216 and r = 0.066, P = 0.484, resp.). SARS-CoV-2-specific IgM levels peaked in forth week after onset of COVID-19 pneumonia, while serum IgG levels increased over time more than 8 weeks (Figure 1C). Finally, IgM and IgG levels were assessed to predict the clinical outcome by ROC curves. The area under the ROC curve (AUC) for IgM antibody was 0.681 (95% confidence interval: 0.517-0.845, P = 0.024), while AUC for IgG antibody was 0.613 (95% confidence interval: 0.443-0.784, P = 0.158) (Figure 1D).

**Figure 1:**
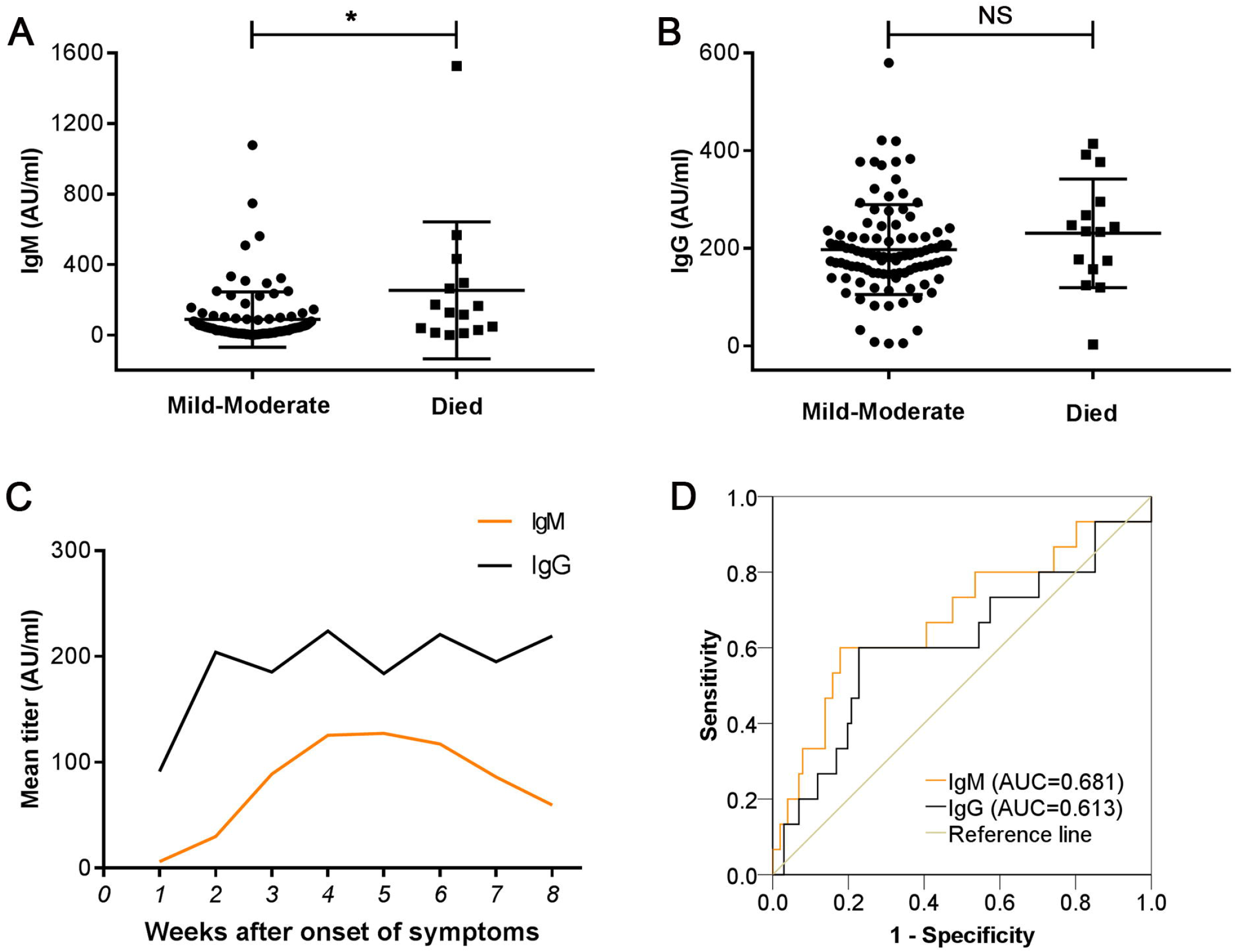
Serum IgM and IgG antibodies in patients with COVID-19 pneumonia. (A) Serum IgM levels were significantly higher in died group. *P < 0 05. (B) No difference found in IgG levels between mild-moderate group and died group. NS: Non-significant. (C) Dynamics of serum IgM and IgG. (D) Accuracy of serum IgM levels in predicting the mortality of patients.

### Case-control study paired by age

As reported, antibodies to coronaviruses are higher in older compared with younger adults^9^. To minimize confounding effects from age, a paired case-control study was further done. Each deceased case was matched with three mild-moderate patients of similar age (age difference within four years). Leucocyte count and neutrophil count of patients on admission were significantly higher in died group than mild-moderate patients (9.4 [IQR 7.3-11.5] vs 6.4 [IQR 4.5-7.1], P = 0.004; 8.2 [IQR 6.2-10.0] vs 4.6 [IQR 2.6-5.7], P < 0.001; Table 3), while lymphocyte count and eosinophil count of patients were significantly lower in died group than mild-moderate patients (0.7 [IQR 0.5-0.9] vs 1.1 [IQR 0.7-1.5], P = 0.017; 0 [IQR 0-0] vs 0.1 [IQR 0-0.1], P = 0.007; Table 3). Blood urea nitrogen and prothrombin time on admission were higher in deceased patients (BUN 10.9 mmol/L [7.5-11.9]; PT 15.5 s [14.6-16.6]) than mild-moderate patients (BUN 5.9 mmol/L [3.8-6.3], P = 0.001; PT 14.5 s [13.6-14.6], P = 0.029). Serum IL6 was higher in deceased patients (15 patients included, 83.4 pg/mL [14.0-65.2]) than mild-moderate patients (41 patients included, 36.7 pg/mL [2.8-55.3], P = 0.032).

**Table 3:** Clinical characters of patients in paired case-control study.

|  | **Total (n=60)** | **Mild-Moderate**<br>**(n=45)** | **Died (n=15)** | **P value** |
| --- | --- | --- | --- | --- |
| Age, years | 68.8 (60.0-78.5) | 68.7(60.0-79.0) | 68.9(57.0-79.0) | 0.948 |
| Men | 37(62%) | 25(56%) | 12(80%) | 0.092 |
| Any comorbidities | 51(85%) | 37(82%) | 14(93%) | 0.297 |
| Positive SARS-CoV-2 RNA | 46(77%) | 34(76%) | 12(80%) | 0.724 |
| Hospital stay | 22.2(17.3-30.0) | 22.0(13.0-30.0) | 22.6(21.0-29.0) | 0.732 |
| Hemoglobin, g/L | 125.2(113.3-137.0) | 122.6(111.5-131.0) | 133.1(114.0-149.0) | 0.132 |
| Alanine aminotransferase, U/L | 30.3(17.0-38.8) | 30.2(14.5-39.0) | 30.5(18.0-39.0) | 0.956 |
| Aspartate aminotransferase, U/L | 33.4(21.0-41.5) | 32.7(21.0-39.5) | 35.3(21.0-45.0) | 0.611 |
| Albumin, g/L | 34.3(29.8-37.4) | 35.1(30.4-38.7) | 31.7(27.4-37.3) | 0.101 |
| Total bilirubin, μmol/L | 11.3(7-13.9) | 10.7(6.5-14.1) | 13.3(9.2-13.9) | 0.105 |
| Lactate dehydrogenase, U/L | 346.5(224.0-295.0) | 328.7(212.0-366.0) | 399.9(262.0-524.0) | 0.275 |
| Blood urea nitrogen, mmol/L | 7.1(4.2-8.3) | 5.9(3.8-6.3) | 10.9(7.5-11.9) | 0.001 |
| Creatinine, μmol/L | 112.2(59.3-98.0) | 96.4(57.5-90.0) | 159.9(80.0-122.0) | 0.158 |
| Prothrombin time, second | 14.8(13.8-15.4) | 14.5(13.6-14.6) | 15.5(14.6-16.6) | 0.029 |
| D-dimer, μg/mL | 3.9(0.4-4.0) | 3.2(0.3-2.6) | 5.9(0.8-6.8) | 0.146 |
| IgM, AU/ml | 119.5(13.2-122.0) | 74.7(10.8-82.0) | 253.8(29.2-296.2) | 0.019 |
| IgG, AU/ml | 208.6(151.2-251.0) | 201.3(149.0-246.9) | 230.7(157.3-295.5) | 0.301 |
| Antibody detection after onset, day | 31.6(26.3-38.8) | 32.0(26.5-40.0) | 30.1(26.0-32.0) | 0.339 |
| Leucocytes, $\times 10^9$ per L | 7.1(4.7-8.4) | 6.4(4.5-7.1) | 9.4(7.3-11.5) | 0.004 |
| Neutrophils, $\times 10^9$ per L | 5.5(2.9-7.0) | 4.6(2.6-5.7) | 8.2(6.2-10.0) | <0.001 |
| Lymphocytes, $\times 10^9$ per L | 1.0(0.6-1.4) | 1.1(0.7-1.5) | 0.7(0.5-0.9) | 0.017 |
| Monocytes, $\times 10^9$ per L | 0.5(0.4-0.6) | 0.5(0.5-0.6) | 0.5(0.3-0.7) | 0.848 |
| Eosinophils, $\times 10^9$ per L | 0.1(0.0-0.1) | 0.1(0.0-0.1) | 0.0(0.0-0.0) | 0.007 |
| Basophils, $\times 10^9$ per L | 0.0(0.0-0.0) | 0.0(0.0-0.0) | 0.0(0.0-0.0) | 0.968 |
| Platelets, $\times 10^9$ per L | 222.3(150.0-300.0) | 225.4(158.5-301.5) | 212.9(113.0-295.0) | 0.267 |

Serum IgM was higher in deceased patients (IgM 253.8 AU/ml [29.2-123.3]) than mild-moderate patients (IgM 74.7 AU/ml [10.8-82.0], P = 0.019), but not IgG (Table 3, Figure 2A). We also explored the correlations between IgM levels and clinical outcome, serum IgG levels, blood counts, or other involved parameters. Similarly, serum IgM levels were positively correlated with IgG levels and leucocyte count (r = 0.279, P = 0.031; r = 0.260, P = 0.045), and negatively correlated with clinical outcome and albumin levels (r = −0.337, P = 0.008; r = −0.265, P = 0.041). Meanwhile, no significant correlations were found between IgM levels and sex, comorbidities, neutrophil count, lymphocyte count, eosinophil count, blood urea nitrogen levels, creatinine levels, PT, D-dimer levels, IL6 levels or duration between onset and antibody detection in patients with COVID-19 pneumonia (r = −0.131, P = 0.319; r = 0.109, P = 0.409; r = 0.254, P = 0.050; r = 0.039, P = 0.769; r = −0.178, P = 0.173; r = −0.026, P = 0.854; r = −0.110, P = 0.404; r = 0.098, P = 0.457; r = 0.127, P = 0.335; r = 0.116, P = 0.394 and r = 0.085, P = 0.521, resp.). Finally, IgM levels were assessed to predict the clinical outcome by ROC curves. The area under the ROC curve (AUC) for IgM levels was 0.704 (95% confidence interval: 0.534-0.873, P = 0.019), while AUC for IgG levels was 0.593 (95% confidence interval: 0.416-0.769, P = 0.286) (Figure 2B).

**Figure 2:**
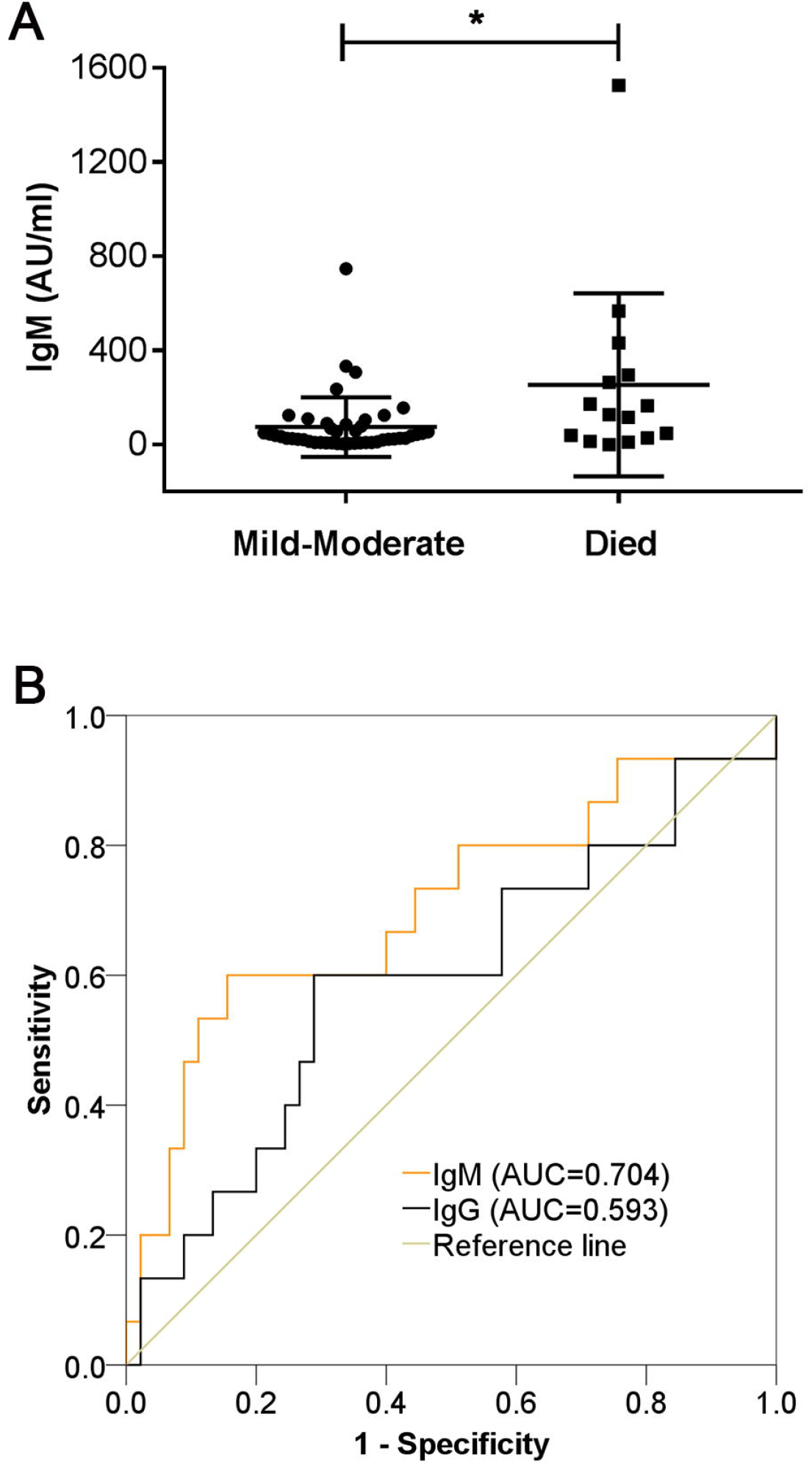
Serum IgM and IgG antibodies in case-control study paired by age. (A) Serum IgM levels were significantly higher in died group in case-control study. (D) Accuracy of serum IgM levels in predicting the mortality of paired patients.

## Discussion

In this retrospective study, we analyzed the levels of serum IgM, IgG and other clinical parameters in 116 hospitalized patients with confirmed COVID-19 pneumonia. The results showed that serum IgM levels were higher in died group, negatively correlated with clinical outcome, and predicted the mortality in patients with COVID-19 pneumonia. Thus, elevated SARS-CoV-2 IgM levels indicate poor outcome.

SARS-CoV-2 is a coronavirus with higher transmission trend and weaker pathogenicity compared with severe acute respiratory syndrome-related coronavirus (SARS-CoV) ^6,10^. Most infected patients show mild symptoms, while some died of fatal pneumonia. Clinical characteristics evaluation, risk factors identification and epidemiological modelization have been well studied, while no effective drugs have been developed ^10-12^. Host immune responses play a key role in virus elimination. Histological examination of lungs from patients who died of COVID-19 revealed bilateral diffuse alveolar damage with cellular exudates, accompanied by interstitial mononuclear inflammatory infiltrates ^13^. Meanwhile, peripheral T cells counts were substantially reduced, while their status was hyperactivated ^13,14^. Raised SARS-CoV-2-specific antibodies have been detected in COVID-19 patients ^7^, but little is known about their dynamics and role in COVID-19 pneumonia. SARS-CoV specific IgG antibodies persisted for a long time over three months, though the SARS-CoV specific IgM peaked in third week and remained measurable for a much shorter period, which were both analyzed by an indirect enzyme-linked immunosorbent assay ^15^. Similarly, as shown in Figure 1C, SARS-CoV-2-specific IgM levels peaked in forth week after onset of COVID-19 pneumonia, then decreased more slowly than SARS-CoV specific IgM, while serum IgG levels increased over 8 weeks. Relatively weaker pathogenicity and prolonged viral shedding provided the rationale for the relatively slower but long-lasting immune response, and provided a strategy of isolation of infected patients and optimal antiviral interventions in the future ^6,11^. The profile of SARS-CoV-2 specific IgM and IgG antibodies might be helpful in the diagnosis and in epidemiologic surveys. Both matched and unmatched analyses yielded similar results that elevated SARS-CoV-2 specific IgM levels indicated poor outcome in patients with COVID-19 pneumonia. Similarly, another team found that a higher titer of antibody titer was associated with a worse clinical classification ^16^. The reason is speculated as follows. Firstly, at least in some stages, high antibody levels indicate high viral load, which is a prognostic factor for poor outcome ^17^. Secondly, a neutralizing antibody can bind to the surface spike protein of coronaviruses and mediate viral entry into IgG Fc receptor-expressing cells, indicating complex roles of antibodies in viral elimination ^18^. Furthermore, the study demonstrates that serum IgM levels were significantly correlated with leucocyte count, indicating more severe inflammation associated with coronavirus infection or secondary infection.

This study also has limitations. It is a single-centre retrospective study with limited cases. Majority of patients admitted in our hospital were critically ill, so population bias exists. In addition, results of viral load and antibody re-testing were not available.

In summary, our results demonstrate that dynamics of IgM and IgG antibodies against SARS-CoV-2 was similar with that of SARS-CoV. Differently, elevated serum IgM levels indicate poor outcome in patients with COVID-19 pneumonia.

## Data Availability

With the permission of the corresponding authors, the raw data without names and identifiers are available on reasonable request. Data can be provided after publication of this study. Once the data are approved to be made public, the researchers will provide an email address for communication. The corresponding authors will make a decision based on the research objectives and plan provided.

## Contributors

ZH, XG and ZC made substantial contributions to the study design. JL and ZW was in charge of the manuscript draft. ZW and HL took responsibility for obtaining ethical approval. CY and ZH took responsibility for data acquisition. CY and JL made main contributions to data analysis and interpretation. JL and HL participated in the diagnosis and treatment of health professionals. ZC and SW made substantial revisions to the manuscript.

## Acknowledgments

This work was supported by National Natural Science Foundation of China (No.81702989)

